# The age paradox in post-infectious sequelae: physiological reserve outweighs chronological age in Long COVID susceptibility

**DOI:** 10.64898/2026.02.24.26346989

**Authors:** Alaleh Azhir, Jingya Cheng, Jiazi Tian, Ingrid V. Bassett, Chirag J. Patel, Jeffrey G. Klann, Shawn N. Murphy, Hossein Estiri

**Affiliations:** Department of Medicine, Massachusetts General Hospital, Boston, MA, USA; Department of Medicine, Brigham and Women’s Hospital, Boston, MA, USA; Department of Biomedical Informatics, Harvard Medical School, Boston, MA, USA; Department of Neurology, Massachusetts General Hospital, Boston, MA, USA

**Keywords:** Post-acute sequelae of SARS-CoV-2, risk factors, Electronic Health Records, Epidemiologic Studies, Biostatistics, SARS-COV-2

## Abstract

**Background:** Older age is widely considered a risk factor for post-acute sequelae of SARS-CoV-2 infection (PASC), typically attributed to immunosenescence and inflammaging. However, whether this association reflects intrinsic biological ageing or accumulated comorbidity burden remains unclear, with implications for clinical risk stratification.

**Methods:** We conducted a retrospective cohort study using the Precision PASC Research Cohort (P2RC) from Mass General Brigham, comprising 133,792 COVID-19 patients from 12 hospitals and 20 community health centres in Massachusetts (March 2020-May 2024). PASC was ascertained using a validated computational phenotyping algorithm. We used generalised estimating equations with cluster-robust variance to model PASC risk, causal mediation analysis to decompose age effects through comorbidity burden and acute severity, and specification curve analysis across 768 analytical specifications to assess robustness.

**Findings:** After adjustment for comorbidity burden, each decade of age was associated with 6% lower odds of PASC (OR 0.94; 95% CI 0.93-0.95). Causal mediation analysis revealed that comorbidities accounted for 145% of the total age effect, indicating inconsistent mediation wherein age’s direct protective effect was masked by its indirect harm through chronic disease accumulation. This protection was age-dependent: adults younger than 65 years retained robust resilience independent of comorbidities (ADE:-0.0042, p<0.001), whereas adults 65 years and older showed complete loss of this protection (ADE: +0.0020, p=0.14).

**Interpretation:** Long COVID susceptibility is driven by physiological reserve rather than chronological age until approximately age 65, beyond which age-related protective mechanisms become exhausted. Risk stratification should prioritise comorbidity burden over birth year in younger adults.

**Funding:** National Institute of Allergy and Infectious Diseases (NIAID).

## Introduction

Older adults bear the greatest burden of acute COVID-19 severity, yet the risk architecture of post-acute sequelae of SARS-CoV-2 infection (PASC) — commonly known as Long COVID — may operate through fundamentally different mechanisms. This distinction matters: if age itself drives Long COVID susceptibility through intrinsic biological processes such as immunosenescence or inflammaging, interventions must target these relatively immutable features of ageing. If, however, chronological age primarily serves as a proxy for accumulated comorbidity burden, the implications for prevention and care shift toward modifiable disease management during acute infections. Indeed, emerging evidence in geroscience increasingly emphasises that biological age, rather than chronological age, more accurately predicts disease outcomes and mortality.[1–3]

PASC encompasses a constellation of symptoms persisting or emerging after the acute phase of COVID-19, including fatigue, cognitive impairment, dyspnoea, and a range of other debilitating effects that significantly diminish quality of life.[4,5] Understanding the factors that govern PASC susceptibility, and critically, disentangling correlated risk factors from causal drivers, is essential for developing targeted interventions.

Current evidence implicates demographic factors, including age, sex, and race, alongside clinical factors such as comorbidity burden and acute infection severity, in determining PASC risk.[6–13] Multiple cohort and longitudinal studies have examined these associations, identifying older age,[14–18] female sex,[6,7,10–12,17] Black race,[6,19] and pre-existing comorbidities[14,17,18,20–22] as consistent correlates. The prevailing interpretation frames older age as an independent risk factor, with proposed mechanisms including immunosenescence, the progressive decline in immune function with age,[24–26] and inflammaging, the chronic, low-grade inflammation characteristic of biological aging.[23,24]

However, prior studies have relied on imprecise case definitions (ICD-10 codes, new-onset diagnoses)[25–32] or subjective ascertainment (survey-based approaches),[33–35] often with limited sample sizes.[26–28,31,33–36] These methodological constraints have led to a systematic underestimation of PASC prevalence, typically identifying only 10–20% of cohorts as affected, [6,7,9,11,13] and have precluded a rigorous examination of whether age operates independently or primarily through its correlation with comorbidity burden. The question is not merely academic: it determines whether clinicians should stratify risk by birth year or by physiological reserve.

In this study, we utilize the Precision PASC Research Cohort (P2RC) (37) to investigate variations in PASC risk across demographic and clinical profiles from 12 hospitals and 20 community health centers. The P2RC, comprising over 81,000 COVID-19 cases with validated PASC ascertainment, provides the statistical power and phenotypic precision necessary to address questions that remained intractable in prior cohort studies. Specifically, we examine whether the apparent age-PASC association persists after rigorous adjustment for comorbidity burden, investigate age-dependent patterns, and characterise the mediation effect among age, disease severity, comorbidity, and PASC.

## Methods

### Study design

We conducted a retrospective cohort study using electronic health records (EHR). The validated P2RC [37] from Mass General Brigham (MGB) includes integrated longitudinal clinical and demographic data from over 130 thousand patients with and without PASC within 12 months of COVID-19 infection from 12 hospitals and 20 community health centres in Massachusetts. The identified conditions were those temporally associated with SARS-CoV-2 infection and not attributable to any pre-existing diagnoses documented in the EHR from 2017 onwards. We do not infer a direct causal relationship between SARS-CoV-2 and these conditions. Instead, we applied an exclusion-based phenotyping approach: for example, a symptom such as shortness of breath was classified as PASC only if it emerged following the index infection and no prior clinical diagnosis (e.g., asthma) was available to account for it. Data was limited to COVID-19 infections between March 6, 2020, and May 8, 2024. Clinical data for follow-up included up to May, 2025. The study is reported in compliance with the Strengthening the Reporting of Observational Studies in Epidemiology (STROBE) guidelines for cohort studies.

### Cohort

We used the P2RC cohort for this study. Post-acute sequelae of SARS-CoV-2 (PASC) in P2RC is defined in accordance with the U.S. National Academies of Sciences, Engineering, and Medicine (NASEM) and the World Health Organisation, in which PASC is an infection-associated chronic condition (IACC) that is unexplained by other conditions and persists for more than two months after the infection episode. [38,39] All patients who tested positive or received a diagnosis code for SARS-CoV-2 infection were included in this study. We monitored the occurrence of SARS-CoV-2 infection from March 2020 to May 2024. Patients who developed PASC within 12 months of infection were designated as the PASC group, while those who did not develop PASC served as the non-PASC group. The study leverages a large sample size, enhancing the robustness and generalisability of the findings. Given that our cohort was derived from a precise validated algorithm, it contains no missing data.

### Ethics

We incorporated demographic characteristics, underlying medical conditions and clinical outcomes into the PASC cohort. This was achieved by linking EHR data from the Enterprise Data Warehouse (EDW) with the Research Patient Data Repository (RPDR). The use of data was approved by the Mass General Brigham (MGB) institutional review board protocol # 2020P001063, with a specified waiver of consent for a data-only retrospective study. Mass General Brigham is an integrated healthcare delivery system serving 1.5 million patients annually.

### Variables

To estimate the risk factors associated with PASC, we evaluated various factors, including vaccination status (Not fully vaccinated and Fully vaccinated), the Charlson Comorbidity Index (calculated with a loyalty score computation[40,41]), and demographic data. Confounders were selected based on a review of the literature, focusing on variables previously associated with both the exposure and the development of PASC. The primary outcome was the presence of PASC, determined by a validated computational algorithm[37] with PASC status being assessed 12 months following each incident infection. Notably, the primary outcome can be recurrent, indicating that an individual may experience multiple episodes of PASC. Similarly, SARS-CoV-2 infections are also repeated measures, reflecting that each individual can develop more than one episode of acute COVID-19. Participants’ information on sex, age, race, and ethnicity was self-reported. Demographic data encompassed age, sex, race (White, Black or African American, Asian, and Other/Unknown), and ethnicity (Hispanic, Non-Hispanic, and Unknown). The severity of infection was classified into 3 levels with increasing severity (non-hospitalisation, hospitalisation only, and ICU or ventilation). Correlation between age and Charlson Comorbidity Index was assessed using Pearson correlation coefficient.

### Primary Analysis

To investigate the risk factors associated with PASC, we applied a generalised estimating equation (GEE)^40^ logistic regression model with an exchangeable working correlation structure. To ensure robust inference, the variance estimator was adjusted to be cluster-robust. We defined the outcome variable P(Y=1) as the occurrence of PASC. The predictor variables included age (modeled as a continuous variable scaled per 10-year increase), sex, race group, time (year–quarter), Charlson Comorbidity Index, Hispanic ethnicity, disease severity, and vaccination status. Linearity of continuous covariates was assessed prior to model fitting. Because the effect of the Charlson Comorbidity Index on the risk of developing PASC did not satisfy the linearity assumption, it was modeled using restricted cubic splines. The model was specified as: logit(P(Y=1)) = β0+ β1 (Age by decade) + β2 (Sex) + β3 (Race) + β4 (year quarter) + β5 (Charlson Index spline) + β6 (Ethnicity) + β7 (Severity) + β8 (Vaccination). This approach allowed us to account for the correlation of multiple infections within the same individual. Then, Odds ratios (ORs) and 95% confidence intervals (CIs) were estimated. All models were adjusted for age, sex (reference: female), race (reference: White), ethnicity (reference: non-Hispanic), Charlson Comorbidity Index, and severity of the acute infection (reference: not severe). A two-sided p value <0.05 was considered statistically significant.

### Casual Mediation Analysis

We conducted the causal mediation analyses to decompose the total effect of age on PASC risk into direct and indirect components using the counterfactual framework for causal inference. Two hypothesized mediators were examined in separate analyses: (1) comorbidity burden, measured by the Charlson Comorbidity Index (categorized as 0, 1, 2, 3, 4, or ≥5), and (2) acute COVID-19 severity. Mediation analysis partitions the total effect of an exposure on an outcome into the Average Causal Mediation Effect (ACME), representing the indirect effect transmitted through the mediator, and the Average Direct Effect (ADE), representing the effect through all other pathways. The total effect equals the sum of ACME and ADE. The proportion mediated was calculated as the ratio of ACME to total effect. Values exceeding 100% indicate inconsistent mediation, wherein direct and indirect effects operate in opposite directions. Given biological evidence for age-related changes in immune function and the clinical relevance of age 65 as a threshold in healthcare policy, we conducted pre-specified mediation analyses stratified by age (<65 years versus ≥65 years).

### Specification Curve Analysis

Finally, we performed a Specification Curve Analysis (SCA) to assess the robustness of our findings by systematically evaluating the impact of various analytical decisions on the results. This method involves identifying a range of reasonable analytical choices, generating all possible combinations of these decisions, and conducting the analysis for each specification. The outcomes are then visualised in a specification curve, which displays the estimated effect sizes across all specifications. By examining the spread and central tendency of the results, we can evaluate the stability and reliability of our findings, ensuring that they are not unduly influenced by specific analytical choices. This approach enhances the rigour and transparency of our analysis.

A logistic regression model was applied to each specification. The outcome variable was PASC status, and the exposure was categorized by age groups in decades. The covariates are Comorbidity, year quarter, and severity. Each specification presents the estimated exposure variable for each possible combination of covariates. We evaluated the effect of age on PASC by sex, race, and three-level age groups (<45 years old, 45-65 years old, and >65 years old). A Specification Curve is presented to show each specification’s estimates, CI, and significance of exposure.

All analyses were conducted using R version 4.3.0 (R Project for Statistical Computing), utilising the geepack, specr, lm4, and forestplot packages. The code for analysis is available at: https://github.com/clai-group/PASC_Risk_study/blob/main/stats_risk_github.R.

## Results

In the MGB P2RC, which is comprised of 133,792 patients diagnosed with 171,050 episodes/incidences of SARS-CoV-2 infection, 24,833 individuals developed post-acute sequelae of SARS-CoV-2 within 12 months of infection, while 108,959 did not exhibit PASC (Table 1). For more information about cohort development and validation, see Azhir et al (2024).^39^

**Table 1.** Summary statistics of the study population.

|  | **Long COVID**<br>**(N=24,833)** | **No Long COVID**<br>**(N=108,959)** | **Overall**<br>**(N=133,792)** |
| --- | --- | --- | --- |
| **Age Mean (SD)** | 60.5 (17.1) | 56.8 (17.1) | 57.5 (17.2) |
| **Sex** |  |  |  |
| F | 16,239 (65.4%) | 68,467 (62.8%) | 84,706 (63.3%) |
| M | 8,594 (34.6%) | 40,492 (37.2%) | 49,086 (36.7%) |
| **Race** |  |  |  |
| White | 18,705 (75.3%) | 82,035 (75.3%) | 100,740 (75.3%) |
| Black | 2,264 (9.1%) | 9,177 (8.4%) | 11,441 (8.6%) |
| Asian | 846 (3.4%) | 4,581 (4.2%) | 5,427 (4.1%) |
| Other | 2,557 (10.3%) | 10,722 (9.8%) | 13,279 (9.9%) |
| Unknown | 461 (1.9%) | 2,444 (2.2%) | 2,905 (2.2%) |
| **Vaccination** |  |  |  |
| Not-vaccinated | 8,769 (35.3%) | 36,595 (33.6%) | 45,364 (33.9%) |
| Vaccinated | 16,064 (64.7%) | 72,364 (66.4%) | 88,428 (66.1%) |
| **Ethnicity** |  |  |  |
| Non Hispanic | 21,603 (87.0%) | 92,937 (85.3%) | 114,540 (85.6%) |
| HISPANIC | 1,268 (5.1%) | 5,886 (5.4%) | 7,154 (5.3%) |
| Unknown | 1,962 (7.9%) | 10,136 (9.3%) | 12,098 (9.0%) |
| **Severity** |  |  |  |
| Non-hospitalized | 21,303 (85.8%) | 101,167 (92.8%) | 122,470 (91.5%) |
| Hospitalized | 2,436 (9.8%) | 6,060 (5.6%) | 8,496 (6.4%) |
| ICU/Ventilation | 1094 (4.4%) | 1,732 (1.6%) | 2,826 (2.1%) |
| **Charlson Index** Mean (SD) | 2.87 (2.53) | 2.06 (2.19) | 2.21 (2.28) |

The mean age of the cohort was 57.5 years. Approximately 63.3 percent of the participants were female, 75.3 percent were White, 8.6 percent were Black or African American, and 5.3 percent were Hispanic.

Among the patients with PASC, the mean age was 60.5 years. Approximately 65.4 percent were female, 75.3 percent were White, 9.1 percent were Black or African American, and 5.1 percent were Hispanic. 6.4% of the cohort was admitted to the hospital, and 2.1% required ICU admission or mechanical ventilation. Among patients with PASC, 64.7% were vaccinated, compared to 66.1% in the overall cohort. The mean Charlson Comorbidity Index was 2.21 in the overall cohort and 2.87 among the patients with PASC.

Age and Charlson Comorbidity Index were strongly correlated (r = 0.698, 95% CI: (0.695, 0.701), P < 0.001; Figure S1). Overall, the likelihood of developing PASC decreased with increasing age (Figure 1). Each ten-year increase in age is associated with a 6 percent decrease in the odds of developing PASC (OR, 0.94; 95% CI, (0.93, 0.95); P < 0.001). Males had a 19 percent decreased likelihood of developing PASC (OR, 0.81; 95% CI, (0.79, 0.84); P < 0.001). Compared to White patients, Black patients had higher odds of developing PASC (OR, 1.06; 95% CI, (1.01, 1.12); P = 0.013). The odds of developing PASC were 9 percent lower for Asian patients compared to White patients (OR, 0.91; 95% CI, (0.85, 0.98); P = 0.014). Vaccinated individuals had a 6 percent lower likelihood of developing PASC (OR, 0.94; 95% CI, (0.91, 0.98); P = 0.0017). Hispanic ethnicity was not significantly associated with PASC (OR, 0.94; 95% CI, (0.88,1.01); P = 0.083). Patients who were hospitalized had 35 percent higher odds of developing PASC (OR, 1.35; 95% CI, (1.29, 1.42); P < 0.001), and those who required ICU admission or mechanical ventilation had 93 percent higher odds of PASC (OR, 1.93; 95% CI, (1.80, 2.07); P < 0.001), compared to non-hospitalized patients.

**Figure 1.**
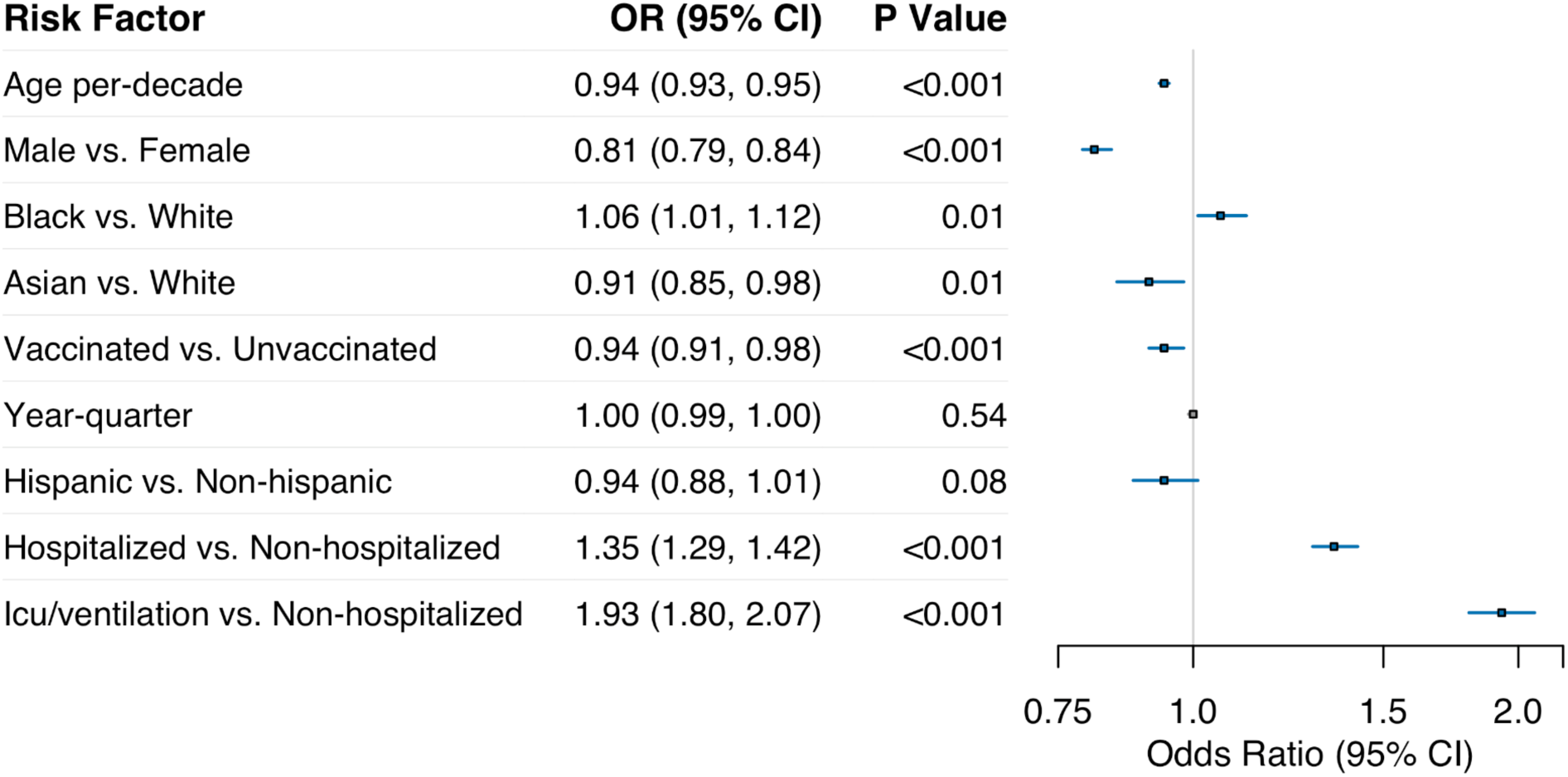
Forest plot of Long COVID across demographic and clinical characteristics in the total study population

The association between the Charlson Comorbidity Index and PASC risk demonstrated a non-linear relationship (Figure 2). Compared to individuals with no comorbidities (Charlson Index = 0), those with a Charlson Index of 5 had 2.73 times the odds of developing PASC (OR = 2.73), while those with a Charlson Index of 10 had 3.48 times the odds (OR = 3.46). The relationship showed a steep initial increase at lower comorbidity levels, with the rate of increase attenuating at higher Charlson Index values.

**Figure 2.**
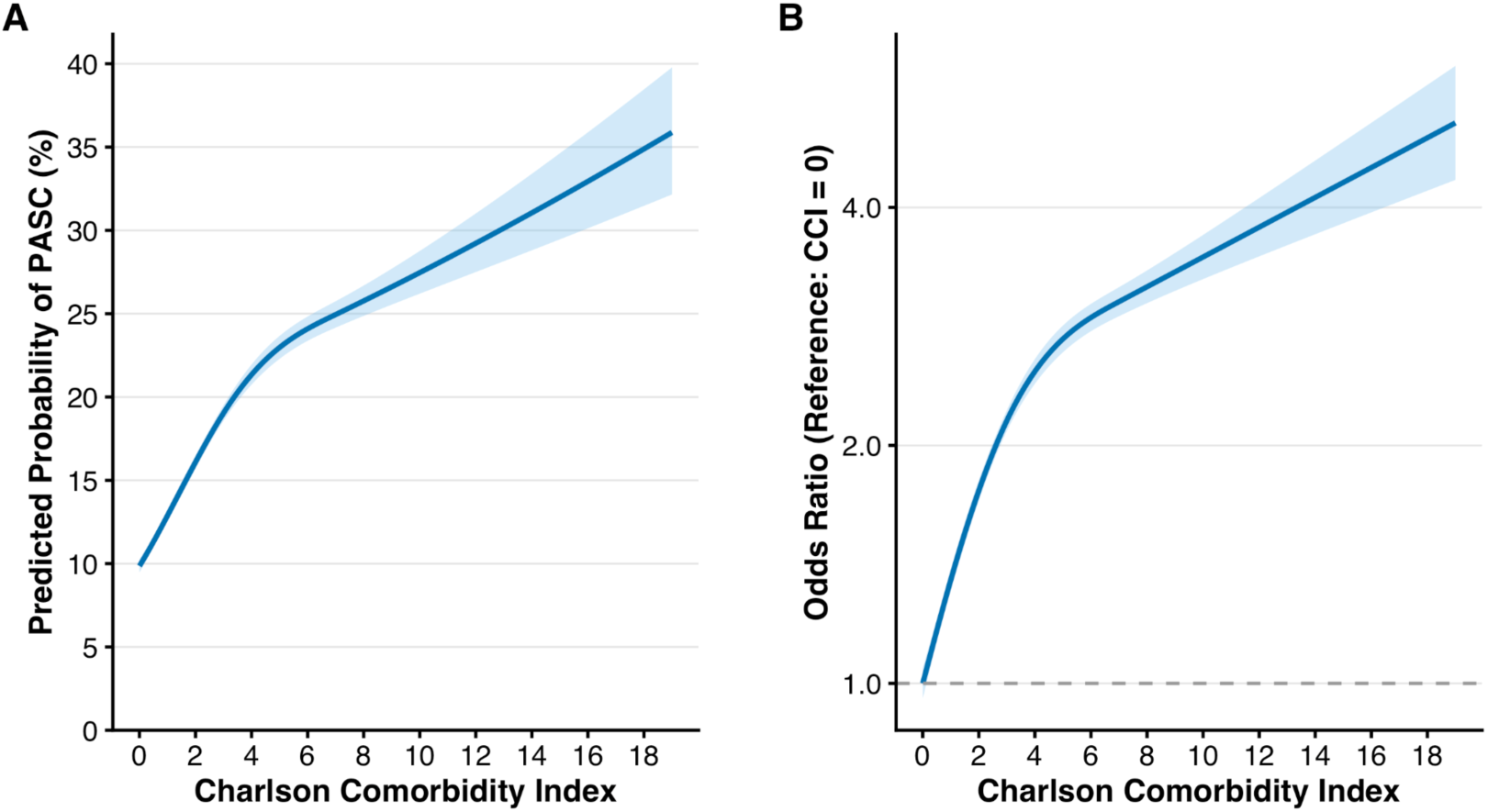
Association Between Charlson Comorbidity Index and PASC Risk

Causal mediation analysis revealed distinct pathways through which age influences PASC risk (Figure 3). Age increased comorbidity burden, which in turn increased PASC risk, yielding a positive indirect effect (ACME: +0.0137 per decade, 95% CI: (0.0127, 0.0145), p<0.001). However, after adjusting for comorbidity burden, age demonstrated a protective direct effect on PASC (ADE: −0.0042, 95% CI: (−0.0055, −0.0032), p<0.001). The total effect of age on PASC was positive (+0.0094, 95% CI: (0.0091, 0.0097), p<0.001), with comorbidities accounting for 145% of this effect. This proportion exceeding 100% indicates inconsistent mediation, wherein the indirect effect through comorbidity accumulation masked age’s protective effect. Age reduced acute COVID-19 severity, which in turn reduced PASC risk, yielding a negative indirect effect (ACME: −0.0011, 95% CI: (−0.0012, −0.0010), p<0.001). The direct effect of age remained protective after adjusting for severity (ADE: −0.0079, 95% CI: (−0.0101, −0.0059), p<0.001). The total effect was protective (−0.0090, 95% CI: (−0.0112, −0.0070), p<0.001), with acute severity mediating 12% of this effect.

**Figure 3.**
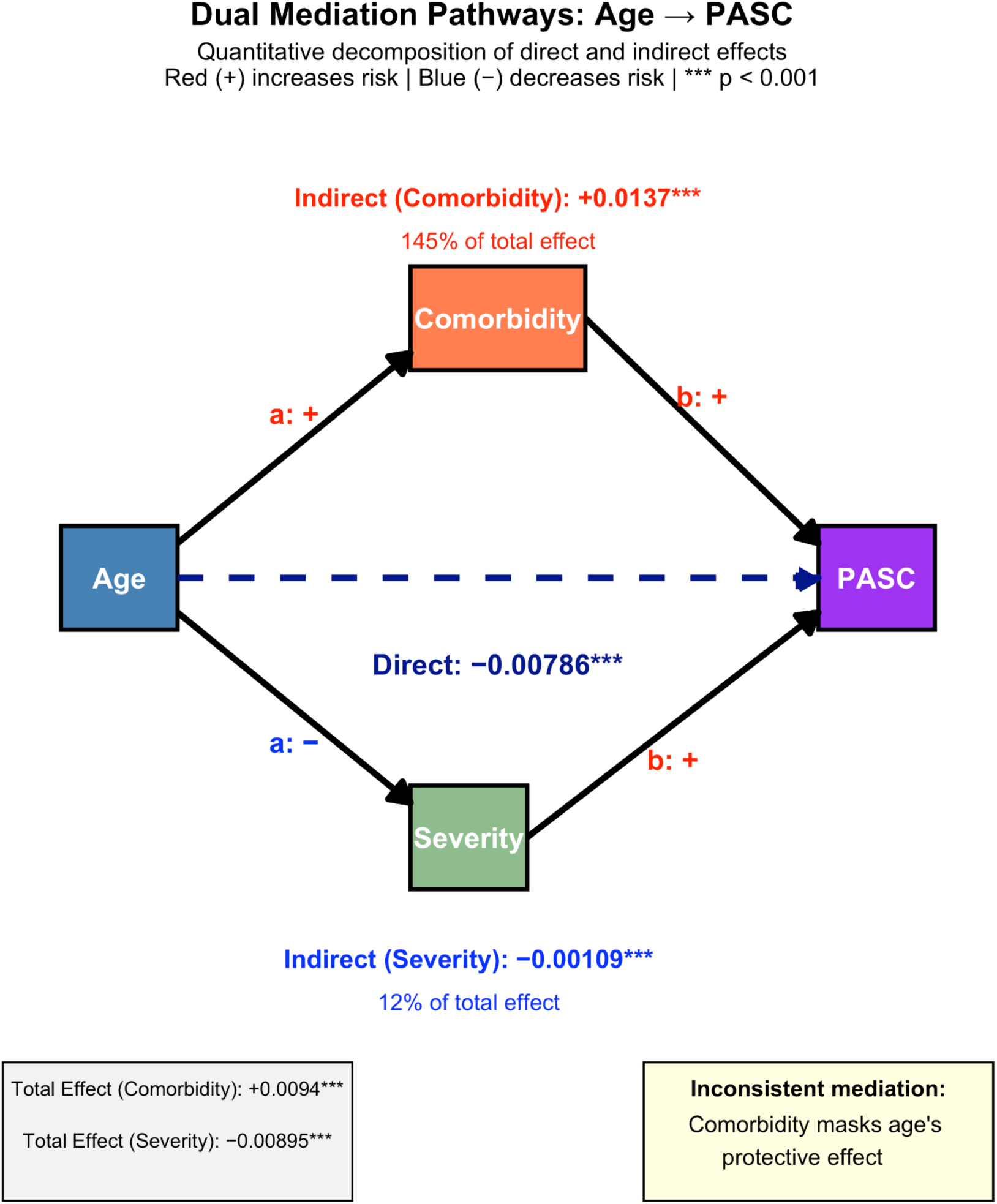
Causal Mediation Analysis of Age Effects on PASC

Age-stratified analyses revealed fundamental differences (Figure S2, Figure S3). In adults <65 years, comorbidity-mediated suppression was pronounced (152% mediated), with preserved direct protective effects (ADE: −0.0042, p<0.001). In adults ≥65 years, age’s protective effect disappeared entirely (ADE: +0.0020, p=0.14), with complete mediation through comorbidity (81%) and severity (26%). Both indirect pathways indicated higher risks of developing PASC in older adults, indicating elimination of age-related resilience beyond age 65.

### Specification Curve Analysis

As previously noted, our investigation revealed a decline in the risk of developing post-acute sequelae of SARS-CoV-2 (PASC) with increasing age. To comprehensively verify this association, we conducted a thorough sensitivity analysis employing Specification Curve Analysis (SCA).

Out of 768 specifications assessed, 384 accounted for comorbidities, with 132 (34.4%) exhibiting negative estimates that were statistically significant, indicative of age acting protectively against PASC, while only 24 (6.25%) specifications with positive estimates were statistically significant (Table S1). The exception was among Black males under 45 years old, who consistently demonstrated positive estimates for age, irrespective of prior comorbidities.

Our findings highlight the intricate role of age and its interaction with comorbidities, particularly represented by the Charlson index. As shown in Figure 4, the regression model’s consideration of comorbidities has a significant impact on the association between age and PASC. Notably, while age emerged as a risk factor for PASC in models that neglected comorbidities, its effect shifted to that of a protective factor when these health conditions were adjusted for.

**Figure 4.**
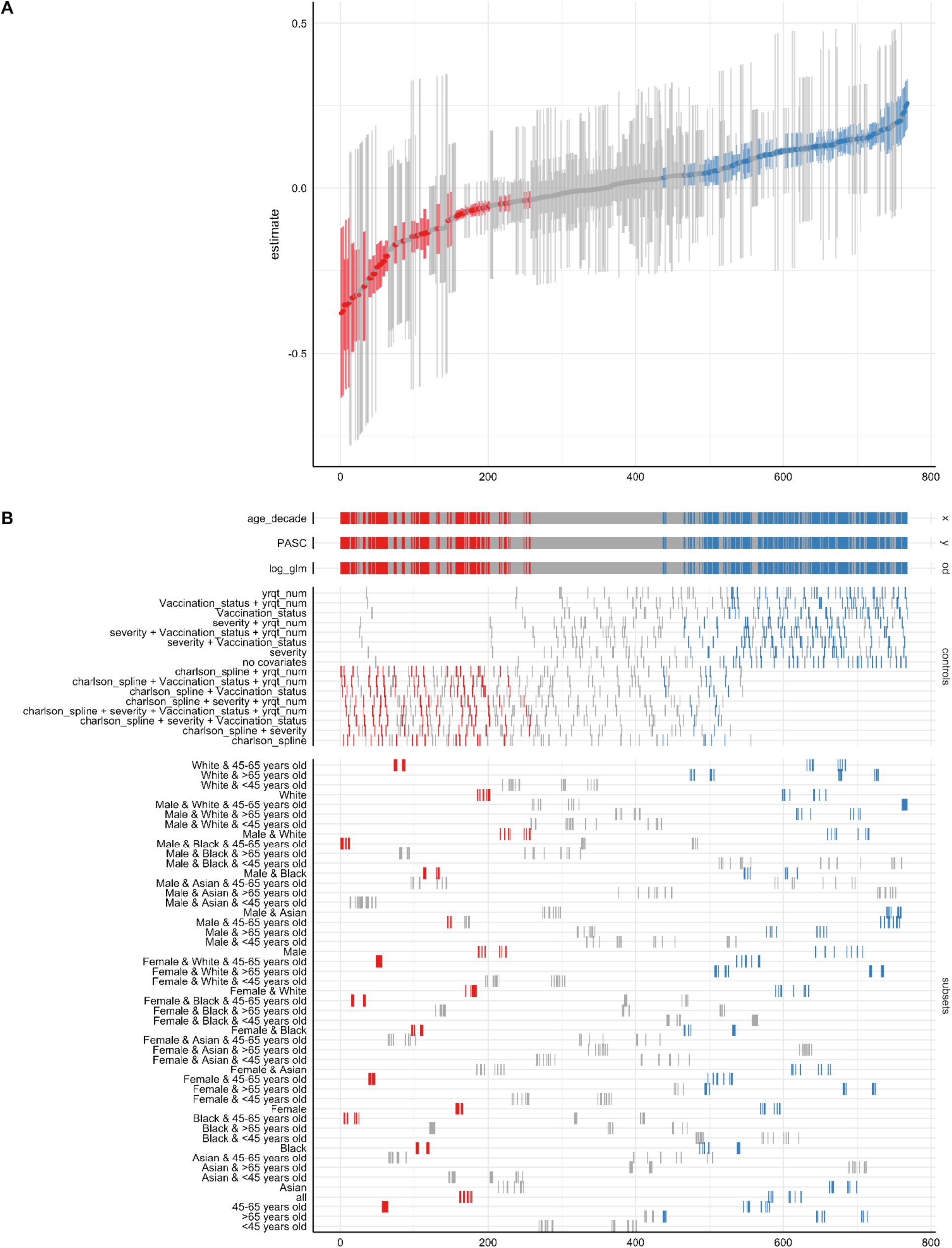
Specification Curve Analysis on the association between age and PASC. A total of 768 specifications were assessed. 70% of specifications that did not include comorbidities exhibited negative estimates for age, indicating that age acts protectively against PASC. Red and blue colours indicate statistically significant estimates, while grey denotes non-significant estimates.

## Discussion

This study challenges the prevailing framework for understanding age-related vulnerability to Long COVID. Using a validated precision cohort of over 130,000 COVID-19 cases, we demonstrate that the apparent association between older age and PASC risk is substantially, and perhaps entirely, mediated by comorbidity burden. When pre-existing conditions are accounted for, age transitions from a risk factor to a protective factor, a finding that remains robust across 768 analytical specifications. This result reframes risk stratification from chronological age toward accumulated disease burden, with implications for both clinical practice and mechanistic understanding.

The dominant explanatory models for age-related PASC susceptibility have invoked immunosenescence and inflammaging, intrinsic features of biological aging that render older adults inherently more vulnerable, regardless of their baseline health status.[23,24,42–44] Our specification curve analysis directly tests this assumption. Among specifications that excluded comorbidity adjustment, age consistently appeared as a risk factor, replicating prior literature.[14–17] However, this association reversed upon adjustment for the Charlson Comorbidity Index, with 34.4% of comorbidity-adjusted specifications showing statistically significant protective effects of age. The implication is stark: studies reporting age as an independent PASC risk factor may have been capturing the shadow of chronic disease rather than ageing biology per se. Our causal mediation analysis provides a formal decomposition of these effects. Age operates through two opposing pathways: a positive indirect effect through comorbidity accumulation (ACME: +0.0137 per decade) and a negative indirect effect through reduced acute infection severity (ACME: −0.0011). The comorbidity pathway dominates, accounting for 145% of the total effect—a proportion exceeding 100% that indicates inconsistent mediation, wherein age’s direct protective effect is masked by its indirect harm through accumulated disease burden. Notably, even after adjusting for both mediators, age retained a modest but statistically significant protective direct effect (ADE: −0.0042 to −0.0079), suggesting that some aspect of ageing beyond comorbidity and acute severity may confer resilience against persistent post-infectious sequelae.

This finding aligns with growing recognition in geroscience that biological age, not chronological age, is the factor most closely associated with disease incidence and outcomes.[1–3] Recent work demonstrates that being phenotypically older than one’s chronological age is more prevalent among those with pre-existing chronic conditions, and this phenotypic age acceleration predicts mortality independent of chronological age.[1,45] Our data suggest that Long COVID susceptibility follows a similar pattern: the physiological reserve depleted by chronic disease, rather than the passage of time itself, determines vulnerability to persistent post-infectious sequelae. The non-linear relationship between comorbidity burden and PASC risk further refines this picture. Compared to individuals without comorbidities, those with a Charlson Index of 5 had 2.73 times the odds of PASC, while those with an index of 10 had only marginally higher odds (OR = 3.48). This attenuation at higher comorbidity levels may reflect survivorship bias; patients with extensive comorbidity burdens who survive acute COVID-19 may represent a selected, physiologically resilient subpopulation, or may indicate that the pathways linking chronic disease to PASC susceptibility become saturated beyond a certain threshold.

We are accustomed to conceptualising older adults as uniformly more vulnerable to disease sequelae, and indeed they often bear the greatest burden of acute illness. But Long COVID appears to operate differently. What matters is not the years accumulated, but the physiological toll those years have exacted; the accumulated burden of chronic conditions that compromise the body’s capacity to resolve infections without lasting consequences. This distinction between chronological age and cumulative physiological burden represents more than semantic refinement; it identifies a modifiable target where none was previously recognised.

However, age-stratified mediation analyses revealed that this protective effect is not uniform across the lifespan. In adults younger than 65 years, the direct protective effect of age remained robust (ADE: −0.0042, p<0.001), with comorbidity burden accounting for 152% of the total effect through inconsistent mediation. In adults 65 years and older, this pattern fundamentally changed: the direct protective effect of age disappeared entirely (ADE: +0.0020, p=0.14), with comorbidity and severity together fully mediating the age-PASC relationship (81% and 26%, respectively). This threshold effect suggests that the physiological mechanisms that confer resilience to Long COVID in younger adults, whether immunological, metabolic, or related to tissue repair capacity, become exhausted or overwhelmed beyond the age of 65. The clinical implication is that risk stratification strategies may need to operate differently across this divide: in younger adults, comorbidity burden should drive clinical concern regardless of age, whereas in older adults, chronological age itself regains prognostic relevance as a marker of diminished reserve.

An alternative, more troubling possibility is that PASC itself induces durable changes in immune function or tissue integrity that increase vulnerability to future episodes. Under this model, each PASC episode would compound risk for subsequent occurrences, suggesting a form of immunological scarring with cumulative consequences. This interpretation is supported by evidence that SARS-CoV-2 reinfection increases the cumulative risk of death, hospitalisation, and sequelae in multiple organ systems,[53,54] with the hazard ratio for at least one sequela increasing from 1.35 after first infection to 2.11 after second infection and 3.00 after three or more infections. While our observational design cannot adjudicate between these mechanisms, the finding underscores that PASC should not be conceptualised as a self-limited condition; for a substantial subset of patients, it may represent the beginning of a chronic, potentially progressive disease trajectory. This observation reinforces the importance of infection prevention strategies, particularly for individuals with a history of prior PASC. Vaccination demonstrated a modest but significant protective effect against PASC (OR 0.94; 95% CI: 0.91–0.98), reducing the risk by 6%. This finding is consistent with emerging evidence suggesting that immunological priming may attenuate post-acute sequelae, even when it does not prevent infection. While the effect size is modest, vaccination represents one of the few modifiable interventions available for PASC risk reduction and should be emphasized in clinical guidance, particularly for patients with elevated comorbidity burdens or prior PASC episodes.

Several additional demographic findings merit discussion. The elevated PASC risk observed in females aligns with broader patterns in autoimmune and infection-associated chronic conditions, where approximately 78% of affected individuals are women.[60,61] This female predominance reflects sex differences in immune regulation, including enhanced antibody production, more robust T helper 2-predominant immune responses, and X chromosome-linked genetic factors that increase susceptibility to chronic inflammatory conditions.[61–63] The female-predominant autoimmune diseases that manifest clinically after acute infections are characterised by persistent inflammation and antibody-mediated pathology,[58, 61] patterns that resemble the immunological signature of Long COVID. The modestly elevated risk among Black individuals, while consistent with prior literature, likely reflects complex interactions between social determinants of health, healthcare access, and unmeasured confounders rather than intrinsic biological differences.

### Limitations

The P2RC relies on structured clinical data, which lacks the depth of information available in clinical narratives and may introduce noise or underreporting. Residual confounding from unmeasured factors, including socioeconomic status, healthcare access, and behavioural variables, cannot be excluded despite our adjustment for available covariates. Causal mediation analysis requires assumptions of no unmeasured confounding of the exposure-mediator, mediator-outcome, and exposure-outcome relationships; violations of these assumptions may bias estimates of direct and indirect effects. Finally, our cohort derives from a single large healthcare system in New England, and generalisability to other populations and healthcare contexts requires validation.

## Conclusions

This study demonstrates that the relationship between age and Long COVID risk is more complex than previously recognised. Rather than reflecting intrinsic biological ageing processes, the elevated PASC rates observed in older adults appear largely attributable to accumulated comorbidity burden, though this protective effect of age diminishes beyond 65 years, where chronological age regains prognostic significance. The non-linear relationship between comorbidity and PASC risk, the opposing mediation pathways through comorbidity burden and acute severity, and the age-dependent loss of resilience collectively argue against uniform risk stratification by birth year. Instead, clinicians should consider physiological reserve as the primary determinant in younger adults while recognising that older adults face compounded vulnerability from both accumulated disease and age-related loss of protective mechanisms. The modest but significant protective effect of vaccination, combined with the substantial recurrence risk in patients with prior PASC, underscores the continued importance of infection prevention across all age groups. As reinfections accumulate in the population, understanding which patients can mount durable recovery and which face progressive post-infectious decline becomes increasingly urgent for clinical care and public health planning.

## Contributors

H.E., J.C., A.A., and I.V.B. conceived, designed, and planned this study. H.E., A.A., J.G.K., J.C., and I.V.B. collected and acquired the data. J.T. performed data preparation and mapping. J.C., and H.E. analysed the data. H.E., and J.C. interpreted the data. J.C., A.A., and H.E. drafted the paper. All authors critically reviewed the paper. J.C., A.A., and H.E. revised the final paper. H.E., J.T., and J.C. had access to all the data in the study. H.E., A.A., J.T., and J.C. verified the underlying data. All authors agreed to submit the manuscript, read and approved the final article.

## Data Sharing Statement

Due to privacy regulations and institutional review board approvals, patient-level data from this study cannot be shared. The computer codes can be accessed at https://github.com/clai-group/PASC_Risk_study.

## Data Availability

Protected Health Information (PHI) restrictions limit the availability of the clinical data, which were used under IRB approval (protocol 2020P001063) for the current study only. As a result, this dataset is not publicly available.

## Acknowledgements

This study has been supported by grants from the National Institute of Allergy and Infectious Diseases (NIAID) R01AI165535.

## References

1. Ho KM, Morgan DJ, Johnstone M, Edibam C. Biological age is superior to chronological age in predicting hospital mortality of the critically ill. Intern Emerg Med. 2023;18: 2019–2028.

2. Wu JW, Yaqub A, Ma Y, Koudstaal W, Hofman A, Ikram MA, et al. Biological age in healthy elderly predicts aging-related diseases including dementia. Sci Rep. 2021;11: 15929.

3. An S, Ahn C, Moon S, Sim EJ, Park S-K. Individualized biological age as a predictor of disease: Korean Genome and Epidemiology Study (KoGES) cohort. J Pers Med. 2022;12: 505.

4. Davis HE, McCorkell L, Vogel JM, Topol EJ. Long COVID: major findings, mechanisms and recommendations. Nat Rev Microbiol. 2023;21: 133–146.

5. Hartung TJ, Neumann C, Bahmer T, Chaplinskaya-Sobol I, Endres M, Geritz J, et al. Fatigue and cognitive impairment after COVID-19: A prospective multicentre study. EClinicalMedicine. 2022;53: 101651.

6. Perlis RH, Santillana M, Ognyanova K, Safarpour A, Lunz Trujillo K, Simonson MD, et al. Prevalence and Correlates of Long COVID Symptoms Among US Adults. JAMA Netw Open. 2022;5: e2238804.

7. Selvakumar J, Havdal LB, Drevvatne M, Brodwall EM, Lund Berven L, Stiansen-Sonerud T, et al. Prevalence and Characteristics Associated With Post-COVID-19 Condition Among Nonhospitalized Adolescents and Young Adults. JAMA Netw Open. 2023;6: e235763.

8. Wang S, Li Y, Yue Y, Yuan C, Kang JH, Chavarro JE, et al. Adherence to Healthy Lifestyle Prior to Infection and Risk of Post-COVID-19 Condition. JAMA Intern Med. 2023;183: 232–241.

9. Richard SA, Pollett SD, Fries AC, Berjohn CM, Maves RC, Lalani T, et al. Persistent COVID-19 Symptoms at 6 Months After Onset and the Role of Vaccination Before or After SARS-CoV-2 Infection. JAMA Netw Open. 2023;6: e2251360.

10. Robineau O, Zins M, Touvier M, Wiernik E, Lemogne C, de Lamballerie X, et al. Long-lasting Symptoms After an Acute COVID-19 Infection and Factors Associated With Their Resolution. JAMA Netw Open. 2022;5: e2240985.

11. Fernández-de-Las-Peñas C, Rodríguez-Jiménez J, Cancela-Cilleruelo I, Guerrero-Peral A, Martín-Guerrero JD, García-Azorín D, et al. Post-COVID-19 Symptoms 2 Years After SARS-CoV-2 Infection Among Hospitalized vs Nonhospitalized Patients. JAMA Netw Open. 2022;5: e2242106.

12. Lam ICH, Wong CKH, Zhang R, Chui CSL, Lai FTT, Li X, et al. Long-term post-acute sequelae of COVID-19 infection: a retrospective, multi-database cohort study in Hong Kong and the UK. EClinicalMedicine. 2023;60: 102000.

13. Morello R, Mariani F, Mastrantoni L, De Rose C, Zampino G, Munblit D, et al. Risk factors for post-COVID-19 condition (Long Covid) in children: a prospective cohort study. EClinicalMedicine. 2023;59: 101961.

14. Bai F, Tomasoni D, Falcinella C, Barbanotti D, Castoldi R, Mulè G, et al. Female gender is associated with long COVID syndrome: a prospective cohort study. Clin Microbiol Infect. 2022;28: 611.e9–611.e16.

15. Bull-Otterson L, Baca S, Saydah S, Boehmer TK, Adjei S, Gray S, et al. Post–COVID Conditions Among Adult COVID-19 Survivors Aged 18–64 and ≥65 Years — United States, March 2020–November 2021. MMWR Surveill Summ. 2022;71: 713.

16. Xie Y, Bowe B, Al-Aly Z. Burdens of post-acute sequelae of COVID-19 by severity of acute infection, demographics and health status. Nat Commun. 2021;12: 6571.

17. Tsampasian V, Elghazaly H, Chattopadhyay R, Debski M, Naing TKP, Garg P, et al. Risk Factors Associated With Post-COVID-19 Condition: A Systematic Review and Meta-analysis. JAMA Intern Med. 2023;183: 566–580.

18. Subramanian A, Nirantharakumar K, Hughes S, Myles P, Williams T, Gokhale KM, et al. Symptoms and risk factors for long COVID in non-hospitalized adults. Nat Med. 2022;28: 1706–1714.

19. Hill EL, Mehta HB, Sharma S, Mane K, Singh SK, Xie C, et al. Risk factors associated with post-acute sequelae of SARS-CoV-2: an N3C and NIH RECOVER study. BMC Public Health. 2023;23: 2103.

20. Thompson EJ, Williams DM, Walker AJ, Mitchell RE, Niedzwiedz CL, Yang TC, et al. Long COVID burden and risk factors in 10 UK longitudinal studies and electronic health records. Nat Commun. 2022;13: 3528.

21. Sudre CH, Murray B, Varsavsky T, Graham MS, Penfold RS, Bowyer RC, et al. Attributes and predictors of long COVID. Nat Med. 2021;27: 626–631.

22. Dzifa Adjaye-Gbewonyo, Anjel Vahratian, Cria G. Perrine, Jeanne Bertolli. Long COVID in Adults: United States, 2022. NCHS Data Brief; 2023 Sep. Report No.: No. 480. doi:10.15620/cdc:132417

23. Franceschi C, Garagnani P, Vitale G, Capri M, Salvioli S. Inflammaging and “Garb-aging.” Trends Endocrinol Metab. 2017;28: 199–212.

24. Franceschi C, Garagnani P, Parini P, Giuliani C, Santoro A. Inflammaging: a new immune–metabolic viewpoint for age-related diseases. Nat Rev Endocrinol. 2018;14: 576–590.

25. Taquet M, Dercon Q, Harrison PJ. Six-month sequelae of post-vaccination SARS-CoV-2 infection: A retrospective cohort study of 10,024 breakthrough infections. Brain Behav Immun. 2022;103: 154–162.

26. Wynberg E, Han AX, Boyd A, van Willigen HDG, Verveen A, Lebbink R, et al. The effect of SARS-CoV-2 vaccination on post-acute sequelae of COVID-19 (PASC): A prospective cohort study. Vaccine. 2022;40: 4424–4431.

27. Kuodi P, Gorelik Y, Zayyad H, Wertheim O, Wiegler KB, Abu Jabal K, et al. Association between BNT162b2 vaccination and reported incidence of post-COVID-19 symptoms: cross-sectional study 2020-21, Israel. NPJ Vaccines. 2022;7: 101.

28. Peghin M, De Martino M, Palese A, Gerussi V, Bontempo G, Graziano E, et al. Post-COVID-19 syndrome and humoral response association after 1 year in vaccinated and unvaccinated patients. Clin Microbiol Infect. 2022;28: 1140–1148.

29. Tannous J, Pan AP, Potter T, Bako AT, Dlouhy K, Drews A, et al. Real-world effectiveness of COVID-19 vaccines and anti-SARS-CoV-2 monoclonal antibodies against postacute sequelae of SARS-CoV-2: analysis of a COVID-19 observational registry for a diverse US metropolitan population. BMJ Open. 2023;13: e067611.

30. Zisis SN, Durieux JC, Mouchati C, Perez JA, McComsey GA. The Protective Effect of Coronavirus Disease 2019 (COVID-19) Vaccination on Postacute Sequelae of COVID-19: A Multicenter Study From a Large National Health Research Network. Open Forum Infect Dis. 2022;9: ofac228.

31. Wisnivesky JP, Govindarajulu U, Bagiella E, Goswami R, Kale M, Campbell KN, et al. Association of Vaccination with the Persistence of Post-COVID Symptoms. J Gen Intern Med. 2022;37: 1748–1753.

32. Yendewa GA, Perez JA, Patil N, McComsey GA. Associations between post-acute sequelae of SARS-CoV-2, COVID-19 vaccination and HIV infection: a United States cohort study. Front Immunol. 2024;15: 1297195.

33. Scherlinger M, Pijnenburg L, Chatelus E, Arnaud L, Gottenberg J-E, Sibilia J, et al. Effect of SARS-CoV-2 Vaccination on Symptoms from Post-Acute Sequelae of COVID-19: Results from the Nationwide VAXILONG Study. Vaccines (Basel). 2021;10. doi:10.3390/vaccines10010046

34. Azzolini E, Levi R, Sarti R, Pozzi C, Mollura M, Mantovani A, et al. Association Between BNT162b2 Vaccination and Long COVID After Infections Not Requiring Hospitalization in Health Care Workers. JAMA. 2022;328: 676–678.

35. Di Fusco M, Sun X, Allen KE, Yehoshua A, Berk A, Alvarez MB, et al. Effectiveness of BNT162b2 BA.4/5 Bivalent COVID-19 Vaccine against Long COVID Symptoms: A US Nationwide Study. Vaccines (Basel). 2024;12. doi:10.3390/vaccines12020183

36. Nehme M, Braillard O, Salamun J, Jacquerioz F, Courvoisier DS, Spechbach H, et al. Symptoms After COVID-19 Vaccination in Patients with Post-Acute Sequelae of SARS-CoV-2. J Gen Intern Med. 2022;37: 1585–1588.

37. Azhir A, Hügel J, Tian J, Cheng J, Bassett IV, Bell DS, et al. Precision phenotyping for curating research cohorts of patients with unexplained post-acute sequelae of COVID-19. Med (N Y). 2024. doi:10.1016/j.medj.2024.10.009

38. National Academies of Sciences Engineering, Medicine. A Long COVID Definition: A Chronic, Systemic Disease State with Profound Consequences. Fineberg HV, Brown L, Worku T, Goldowitz I, editors. Washington, DC: The National Academies Press; 2024.

39. Soriano JB, Murthy S, Marshall JC, Relan P, Diaz JV, WHO Clinical Case Definition Working Group on Post-COVID-19 Condition. A clinical case definition of post-COVID-19 condition by a Delphi consensus. Lancet Infect Dis. 2022;22: e102–e107.

40. loyalty_cohort: Uses heuristics to find patients who get the majority of their care from your hospital system. Github; Available: https://github.com/i2b2plugins/loyalty_cohort

41. Klann JG, Henderson DW, Morris M, Estiri H, Weber GM, Visweswaran S, et al. A broadly applicable approach to enrich electronic-health-record cohorts by identifying patients with complete data: a multisite evaluation. J Am Med Inform Assoc. 2023. doi:10.1093/jamia/ocad166

42. Ramos Jesus F, Correia Passos F, Miranda Lopes Falcão M, Vincenzo Sarno Filho M, Neves da Silva IL, Santiago Moraes AC, et al. Immunosenescence and Inflammation in Chronic Obstructive Pulmonary Disease: A Systematic Review. J Clin Med Res. 2024;13. doi:10.3390/jcm13123449

43. Pawelec G, Adibzadeh M, Pohla H, Schaudt K. Immunosenescence: ageing of the immune system. Immunol Today. 1995;16: 420–422.

44. Gruver AL, Hudson LL, Sempowski GD. Immunosenescence of ageing. J Pathol. 2007;211: 144–156.

45. Levine ME, Lu AT, Quach A, Chen BH, Assimes TL, Bandinelli S, et al. An epigenetic biomarker of aging for lifespan and healthspan. Aging (Albany NY). 2018;10: 573–591.

46. Yousefzadeh MJ, Flores RR, Zhu Y, Schmiechen ZC, Brooks RW, Trussoni CE, et al. An aged immune system drives senescence and ageing of solid organs. Nature. 2021;594: 100–105.

47. Desdín-Micó G, Soto-Heredero G, Aranda JF, Oller J, Carrasco E, Gabandé-Rodríguez E, et al. T cells with dysfunctional mitochondria induce multimorbidity and premature senescence. Science. 2020;368: 1371–1376.

48. Nikolich-Žugich J. The twilight of immunity: emerging concepts in aging of the immune system. Nat Immunol. 2018;19: 10–19.

49. Phetsouphanh C, Darley DR, Wilson DB, Howe A, Munier CML, Patel SK, et al. Immunological dysfunction persists for 8 months following initial mild-to-moderate SARS-CoV-2 infection. Nat Immunol. 2022;23: 210–216.

50. Cheon IS, Li C, Son YM, Goplen NP, Wu Y, Cassmann T, et al. Immune signatures underlying post-acute COVID-19 lung sequelae. Sci Immunol. 2021;6: eabk1741.

51. Cervia-Hasler C, Brüningk SC, Hoch T, Fan B, Muzio G, Thompson RC, et al. Persistent complement dysregulation with signs of thromboinflammation in active Long Covid. Science. 2024;383: eadg7942.

52. Haunhorst S, Bloch W, Wagner H, Ellert C, Krüger K, Vilser DC, et al. Long COVID: a narrative review of the clinical aftermaths of COVID-19 with a focus on the putative pathophysiology and aspects of physical activity. Oxf Open Immunol. 2022;3: iqac006.

53. Bowe B, Xie Y, Al-Aly Z. Acute and postacute sequelae associated with SARS-CoV-2 reinfection. Nat Med. 2022;28: 2398–2405.

54. Al-Aly Z, Bowe B, Xie Y. Long COVID after breakthrough SARS-CoV-2 infection. Nat Med. 2022;28: 1461–1467.

55. Antonelli M, Pujol JC, Spector TD, Ourselin S, Steves CJ. Risk of long COVID associated with delta versus omicron variants of SARS-CoV-2. Lancet. 2022;399: 2263–2264.

56. Xie Y, Choi T, Al-Aly Z. Risk of death in patients hospitalized for COVID-19 vs seasonal influenza in fall-winter 2022-2023. JAMA. 2023;329: 1697–1699.

57. Magnusson K, Kristoffersen DT, Dell’Isola A, Kiadaliri A, Turkiewicz A, Runhaar J, et al. Post-covid medical complaints following infection with SARS-CoV-2 Omicron vs Delta variants. Nat Commun. 2022;13: 7363.

58. Madhi SA, Ihekweazu C, Rees H, Pollard AJ. Decoupling of omicron variant infections and severe COVID-19. Lancet. 2022;399: 1047–1048.

59. Hui KPY, Ho JCW, Cheung M-C, Ng K-C, Ching RHH, Lai K-L, et al. SARS-CoV-2 Omicron variant replication in human bronchus and lung ex vivo. Nature. 2022;603: 715–720.

60. Fairweather D, Frisancho-Kiss S, Rose NR. Sex differences in autoimmune disease from a pathological perspective. Am J Pathol. 2008;173: 600–609.

61. Angum F, Khan T, Kaler J, Siddiqui L, Hussain A. The prevalence of autoimmune disorders in women: A narrative review. Cureus. 2020;12: e8094.

62. Rubtsova K, Marrack P, Rubtsov AV. Sexual dimorphism in autoimmunity. J Clin Invest. 2015;125: 2187–2193.

63. Dou DR, Zhao Y, Belk JA, Zhao Y, Casey KM, Chen DC, et al. Xist ribonucleoproteins promote female sex-biased autoimmunity. Cell. 2024;187: 733–749.e16.

