## Supplementary material for "The age paradox in post-infectious sequelae: physiological reserve outweighs chronological age in Long COVID susceptibility": Table S1, Figure S1, Figure S2, Figure S3

**Table S1.** Specification Curve Analysis outputs summary

|  | Overall | Positive estimate* | Negative estimates* | Insignificant |
| --- | --- | --- | --- | --- |
| With comorbidities | 384 | 24 (6.25%) | 132 (34.4%) | 228 (59.4%) |
| Without comorbidities | 384 | 192 (50%) | 0 | 192 (50%) |
| Black Male under 45 years old with comorbidities | 8 | 8 (100%) | 0 | 8 (100%) |
| Black Male under 45 years old without comorbidities | 8 | 8 (100%) | 0 | 8 (100%) |

### * Only specifications with statistically significant positive/negative estimates are counted.

**Figure S1**. Correlation Between Age and Charlson Comorbidity Index.

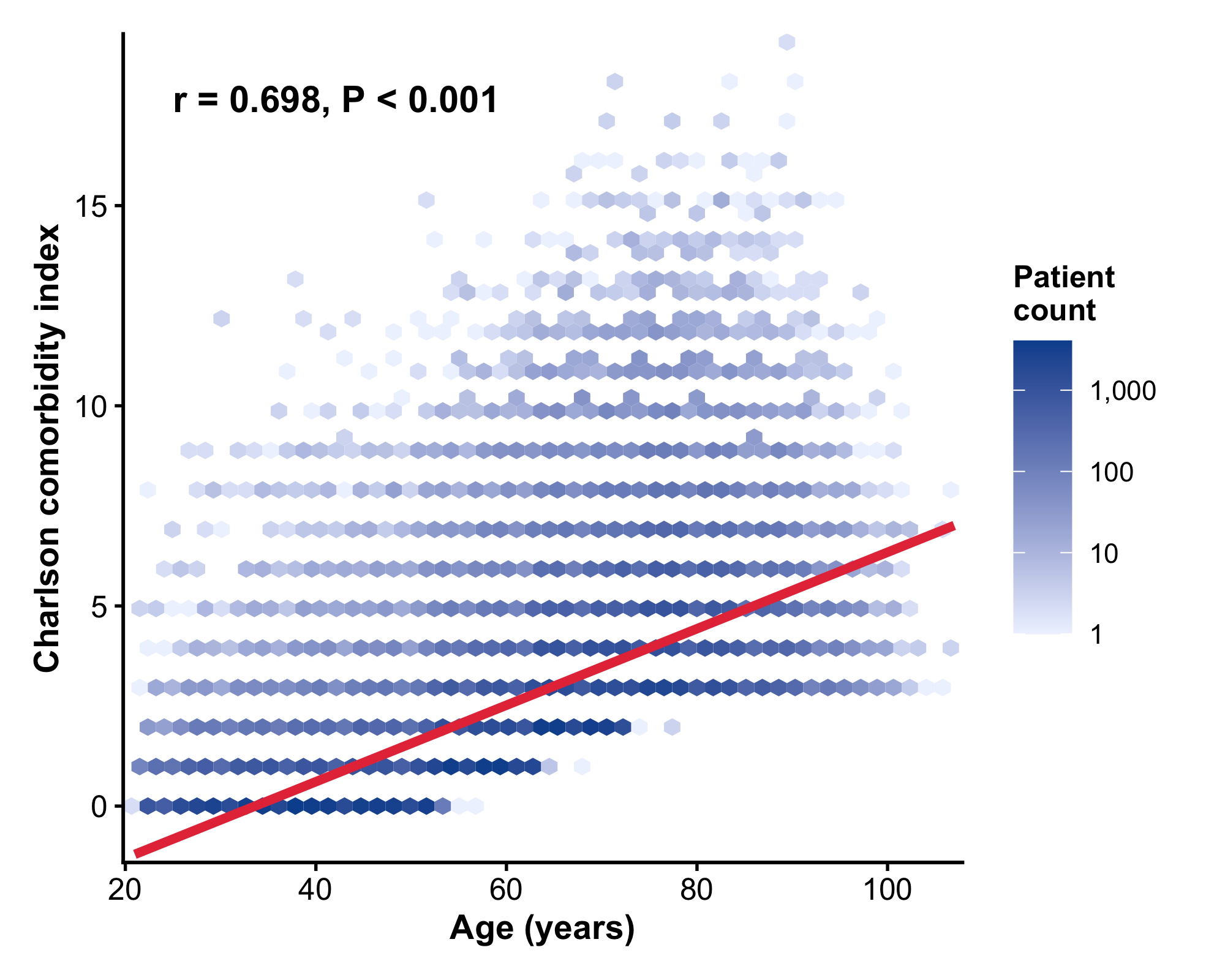

**Figure S2**. Causal Mediation Analysis of Age Effects on PASC in younger adults (age <65)
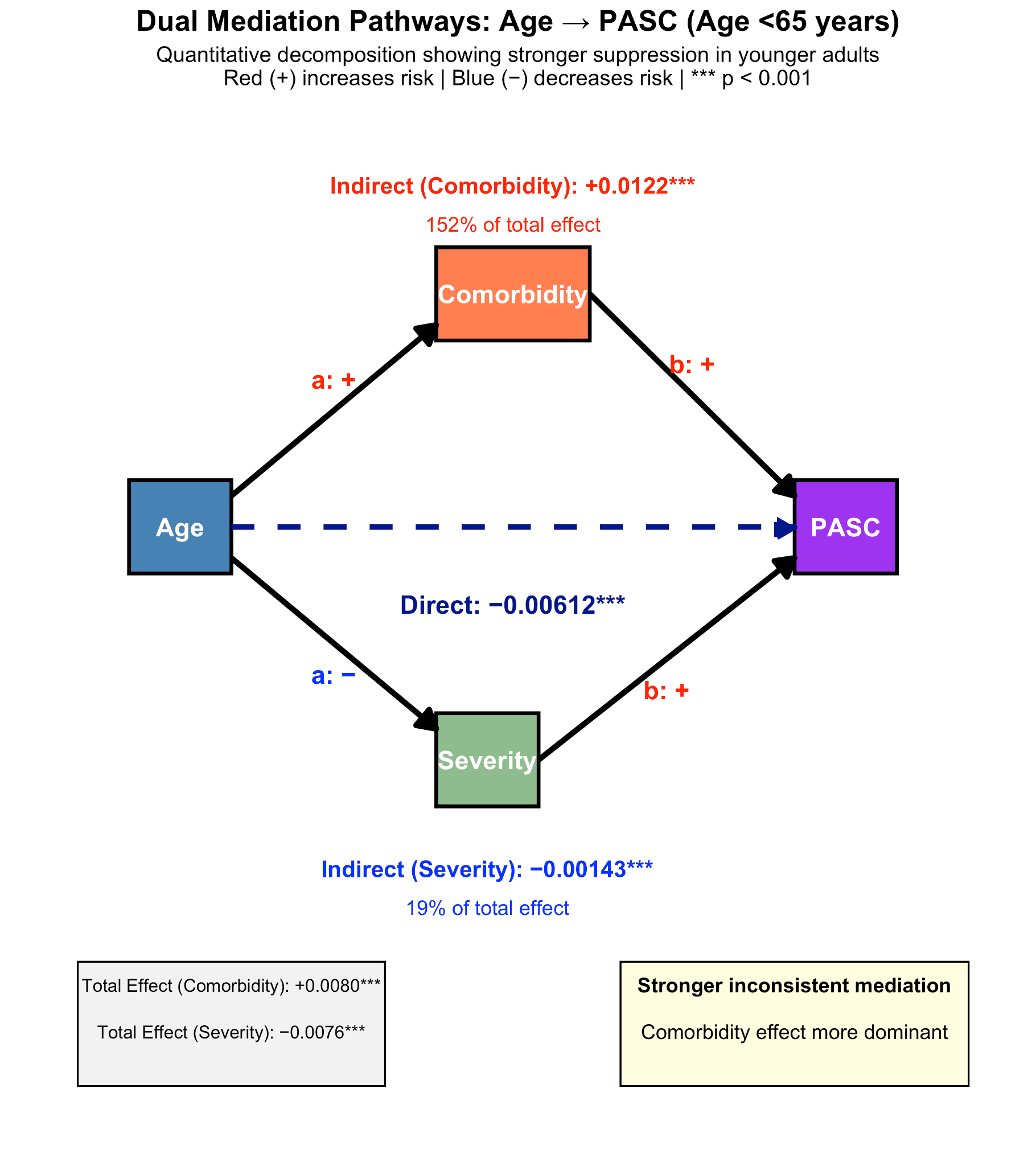

**Figure S3**. Causal Mediation Analysis of Age Effects on PASC in younger adults (age >=65)

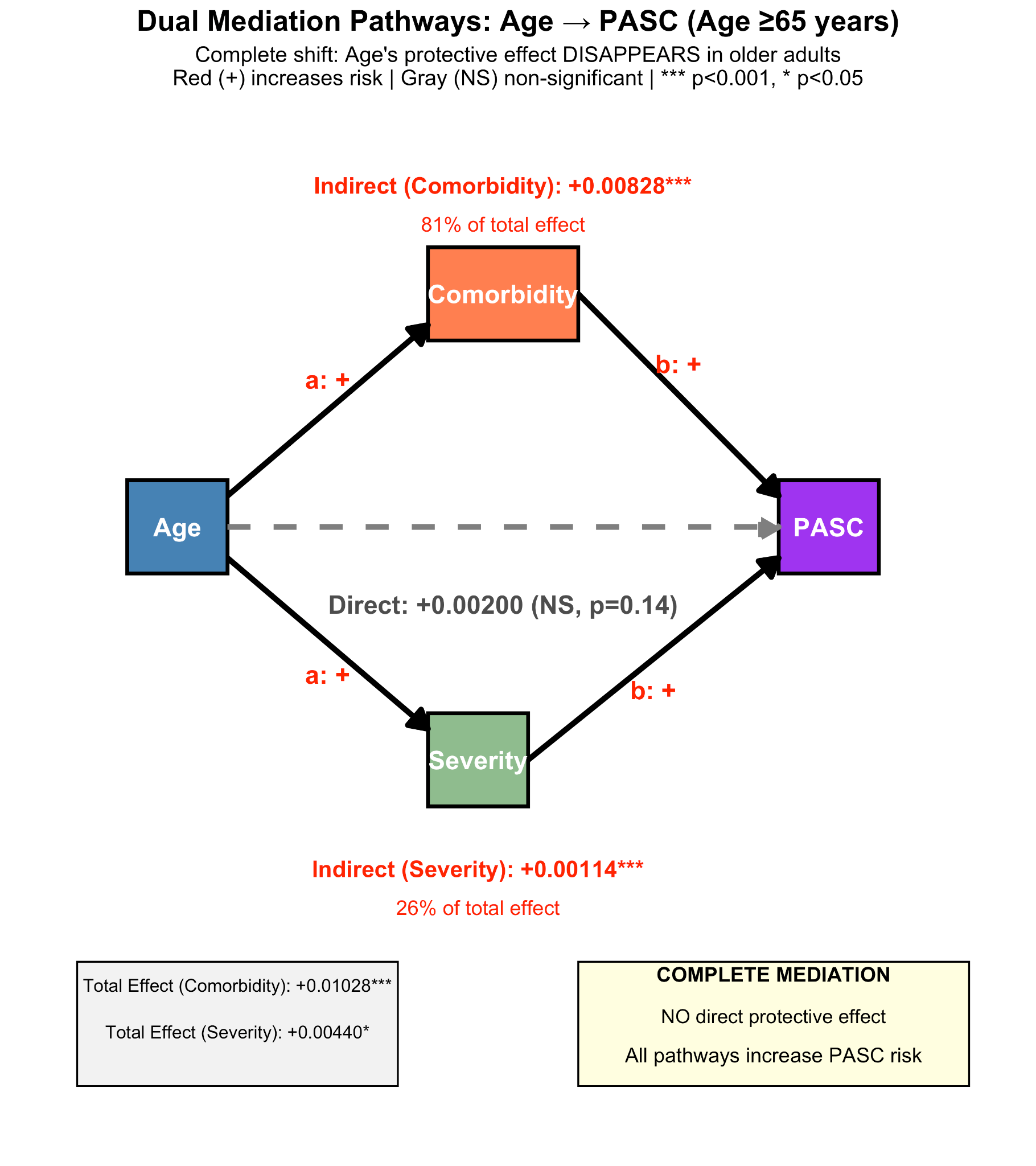
